# Brain morphometrics correlations with age among 352 participants imaged with both 3T and 7T MRI: 7T improves statistical power and reduces required sample size

**DOI:** 10.1101/2024.10.28.24316292

**Authors:** Cong Chu, Tales Santini, Jr-Jiun Liou, Ann D. Cohen, Pauline M. Maki, Anna L. Marsland, Rebecca C. Thurston, Peter J. Gianaros, Tamer S. Ibrahim

## Abstract

**Introduction:** Magnetic resonance imaging (MRI) at 7 Telsa (7T) has superior signal-to-noise ratio to 3 Telsa (3T) but also presents higher signal inhomogeneities and geometric distortions. A key knowledge gap is to robustly investigate the sensitivity and accuracy of 3T and 7T MRI in assessing brain morphometrics. This study aims to (a) aggregate a large number of paired 3T and 7T scans to evaluate their differences in quantitative brain morphological assessment using a widely available brain segmentation tool, FreeSurfer, as well as to (b) examine the impact of normalization methods for subject variability and smaller sample sizes on data analysis.

**Methods:** A total of 452 healthy participants aged 29 to 68 were imaged at both 3T and 7T. Structural T1-weighted magnetization-prepared rapid gradient-echo (MPRAGE) images were processed and segmented using FreeSurfer. To account for head size variability, the brain volumes underwent intracranial volume (ICV) correction using the Residual (regression model) and Proportional (simple division to ICV) methods. The resulting volumes and thicknesses were correlated with age using Pearson correlation and false discovery rate correction. The correlations were also calculated in increasing sample size from 3 to the whole sample to estimate the sample size required to detect aging-related brain variation.

**Results:** 352 subjects (210 females) passed the image quality control with 100 subjects excluded due to excessive motion artifacts on 3T, 7T, or both. 7T MRI showed an overall stronger correlation between morphometrics and age and a larger number of significantly correlated brain volumes and cortical thicknesses. While the ICV is consistent between both field strengths, the Residual normalization method shows markedly higher correlation with age for 3T when compared with the Proportional normalization method. The 7T results are consistent regardless of the normalization method used.

**Conclusion:** In a large cohort of healthy participants with paired 3T and 7T scans, we compared the statistical performance in assessing age-related brain morphological changes. Our study reaffirmed the inverse correlation between brain volumes and cortical thicknesses and age and highlighted varying correlations in different brain regions and normalization methods at 3T and 7T. 7T imaging significantly improves statistical power and thus reduces required sample size.

**Key points:**

1. Compared to 3T, 7T has stronger inverse correlations of total grey matter, subcortical grey matter, and white matter volumes, and mean cortical thickness with age.
2. Compared to 3T, 7T shows a greater number of brain volumes and cortical thicknesses that have statistically significant correlations with age.
3. For comparable statistical power at 3T, the required sample size for 7T is reduced for cortical and subcortical volumes, and substantially reduced for cortical thicknesses.

## Introduction

Magnetic Resonance Imaging (MRI) provides optimal *in vivo* soft tissue contrast and is the method of choice to investigate many cerebral abnormalities such as tumors, atrophy, vascular diseases, demyelinating diseases, trauma, infection, and developmental anomalies (Barisano et al., 2019). The current state of the art clinical usage of MRI could shift from scanners with a static magnetic field of 3 Tesla (T) to the recently FDA-cleared 7T MRI (US FDA, 2024). However, this change is not merely a rescaling of the system but a major engineering challenge.

The 7T MRI offers a higher signal-to-noise ratio (SNR) due to its inherently higher spin signal, as well as improved tissue contrast due to longer T1 and shorter T2 and T2* relaxation times, which help to enhance image and angiography contrasts (Okada et al., 2022; Perera Molligoda Arachchige & Garner, 2023). Moreover, its higher sensitivity to susceptibility differences enhances BOLD contrast (Okada et al., 2022; Perera Molligoda Arachchige & Garner, 2023), which is often used in functional imaging. However, the shorter wavelength of the electromagnetic excitation at 7T increases image inhomogeneity (Ibrahim et al., 2007) and average/local power deposition (Ibrahim & Tang, 2007) for neuroimaging. Both increased average and local power deposition limits the maximum allowed power to be used during the scans (Fiedler et al., 2018). Moreover, the increased sensitivity to susceptibility can cause distortions and artifacts in regions with thin air-brain interfaces, such as the sinus (Truong et al., 2006), depending on the subject’s anatomy. Structural images can also be contaminated with angiography signals, making it difficult for automated segmentation tools to determine brain volumes in specific regions (Choi et al., 2020; Viviani et al., 2017)

Given these trade-offs, there is a need to investigate the sensitivity and specificity of using 3T and 7T MRI to brain morphometrics in a large data set where the same subjects undergo scans at both field strengths. With no clear ground truth in whole brain morphometrics, prior studies have shown that after the human brain reaches its maximum volume between the ages of 25-30 (Fjell et al., 2014; Fjell, 2010), a subject-specific loss of cerebral volume is expected over time. Therefore, MR studies have typically investigated the morphological characteristics and atrophy of brain regions in relation to aging. Studies have found that total grey matter volumes decrease consistently over age, while individual regions showed specificity in their rate of decrease (Fjell et al., 2014; Fjell, 2010). Cortical thicknesses have also been observed to negatively correlate with age (Fjell et al., 2014; Fjell, 2010). White matter volume, on the other hand, differed from grey matter such that it shows modest changes until 40-50 years before a rapid decrease in volume (Fjell et al., 2014; Fjell, 2010).

Compared to 3T, 7T MRI has been shown to provide improved spatial resolution for the same acquisition times (Okada et al., 2022; Perera Molligoda Arachchige & Garner, 2023). However, due to field inhomogeneities, 7T structural images provided restricted performance improvement regarding clinical diagnosis (Springer et al., 2016) and morphometric assessment (Lusebrink et al., 2013; Seiger et al., 2015). These studies, however, were limited by their small sample size of paired 3T and 7T images and by the hardware limitations such as commercial radiofrequency (RF) coils. Our study aims to analyze a large number of same-subject 3T and 7T scans (> 400) to investigate their performance difference in quantitative brain morphological assessment using a widely available brain segmentation tool, FreeSurfer, in addition to homogeneous RF coils that largely eliminate field inhomogeneities and signal voids at 7T (Andrea N Sajewski, 2023; Kim et al., 2016; Krishnamurthy et al., 2019; Santini et al., 2021; Santini et al., 2018). Specifically, we investigated how different statistical analysis and normalization methods may affect the resulting statistical power including correlation strength and sample size. Finaly, provide regression models of the expected regional brain volumes and cortical thicknesses by age.

## Methods

### Participants

The dataset was pooled from multiple studies (NIH RF1AG053504, R01AG053504, P01HL040962, and R01DK110041) recruiting healthy participants under the Institutional Review Board of The University of Pittsburgh, Pittsburgh, USA. Prior to their initial visit, participants underwent a comprehensive informed consent process, which included a detailed review of the study’s objectives. Participants were eligible if they were between the ages of 18 and 80 and had no contraindication to an MRI scan. Additionally, screening for pre-existing dementia was conducted using both the Informant Questionnaire on Cognitive Decline in the Elderly and the Clinical Dementia Rating scale for exclusions.

### Data Collection

Before quality control, in total, 452 subjects had completed paired 3T and 7T MPRAGE sequence with an average (SD) interval of 4.96 (4.16) year. The 7T scans were acquired using a 7T Magnetom system in the sTx mode (single channel) with either the first (16 - combined into one - transmit and 32 receive channels) or second (60 transmit - combined into one - transmit and 32 receive channel) generation of in-house-designed Tic-Tac-Toe RF coil systems. These RF coil systems are known for producing homogeneous images (Andrea N Sajewski, 2023; Kim et al., 2016; Krishnamurthy et al., 2019; Santini et al., 2021; Santini et al., 2018). The 3T scans were acquired with either a Trio or PRISMA systems and utilized an integrated whole-body RF coil for excitation and a commercial 32-ch coil for reception. Description of the acquisition parameters and type of sequences are provided in **Table 1**.

**Table 1.**
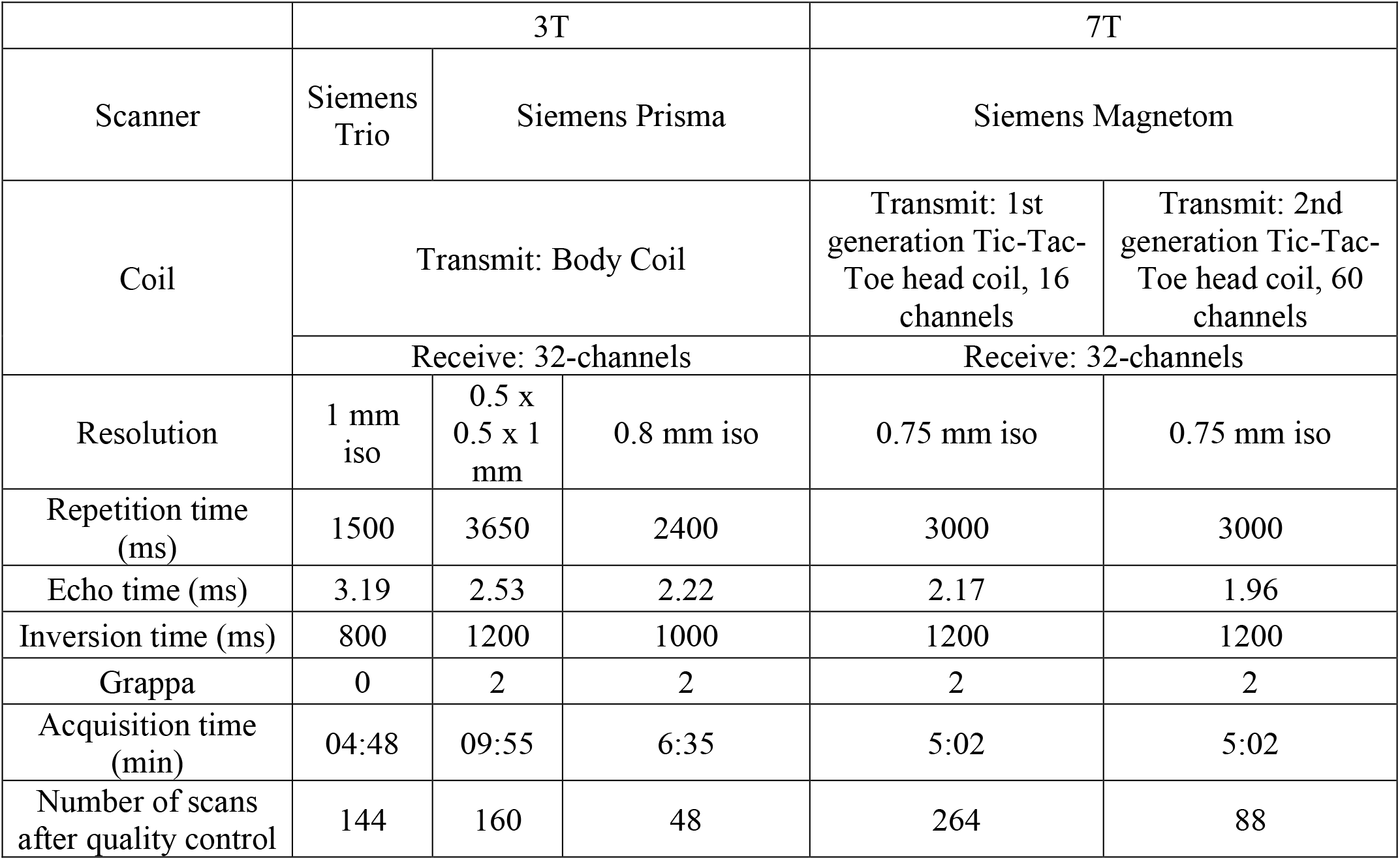
T1-weighted Magnetization Prepared RApid Gradient Echo (MPRAGE) sequence parameters at 3T and 7T.

### Image Processing

Both 3T and 7T scans were processed using the same pipeline. The images were corrected for gradient distortion [https://github.com/Washington-University/gradunwarp] and then underwent intensity bias correction using SPM12 (*Statistical Parametric Mapping: The Analysis of Functional Brain Images*, 2006). Brain stripping was performed using SynthStrip (Hoopes et al., 2022) followed by a 6 DOF rigid registration to MNI space with their respective resolution using Greedy (https://github.com/pyushkevich/greedy). Finally, brain volumes and cortical thicknesses were extracted using FreeSurfer version 7.1.1 (Fischl, 2012) and using the “highres” flag. Intracranial volumes were calculated from the brain mask output of SynthStrip.

The quality control process of the FreeSurfer segmentation output began with classifying the scans into three grades: 1) pass, 2) re-run, and 3) fail. For grade 2 scans, control points were placed on white matter regions that failed to be identified by FreeSurfer. After the re-run with control points, scans were reclassified into either pass of fail. If a subject had either 3T or 7T segmentation classified as failed, both 3T and 7T scans were excluded from the analysis. We also identified and excluded regions that are not consistently segmented due to the presence of dura, or due to the presence of arteries on the 7T images but not on the 3T images. Both issues impact the accuracy as well as the consistency of the FreeSurfer segmentation of the excluded cortical regions on both the 3T and 7T images.

### Statistical Analysis

The volume of the brain regions underwent intracranial volume (ICV) correction using two methods (Wang et al., 2024): In the Residual method, the regions along with ICV and sex were entered into a multiple regression. We extracted the residuals which represented the morphometric information without the effect of ICV and sex. In the Proportional method: the volume regions were divided by their respective ICV and then corrected for sex using regression. Cortical thicknesses were only corrected by sex.

Each region was then correlated with age using Pearson correlations in MATLAB (version R2022a) (MathWorks, Natick MA). Multiple comparison corrections using Benjamini– Hochberg method for False Discovery Rate (FDR) (Benjamini & Hochberg, 1995) were then performed on the p-values within groups separated by cortical volumes, cortical thicknesses, and subcortical volumes. Regions with FDR corrected p-values lower than 10% FDR threshold were considered significantly correlated with age. Linear regression was used to calculate the slope of the correlation. The regions were also fitted with a second order polynomial to estimate the effect of aging on the rate of volume or thickness change. When comparing the correlation coefficient between 3T and 7T data, z-test was performed on the R values undergoing Fisher’s z transformation. The correlation coefficient was statistically stronger than one another when the resulting one-tailed p value was less than 0.05.

To evaluate the effects of the sample size in the number of regions significantly correlated with age, the correlations were calculated in increasing sample size (n = 3 to full sample) for 3T and 7T scans. Each subsample was randomly selected 1000 times without repeating to estimate the error range.

For cortical grey matter volume regions, we also calculated the annual rate of change. The linear regression equation was used to calculate the volume at the median age of the population and the change of volume in one year.

## Results

### Demographics and quality control outcomes

352 subjects out of 452 (female = 210) ranged between 29 and 68 years passed the quality control and were included in the analysis. 100 subjects were excluded due to motion artifacts. Demographics, including sex, race, and years of education, as well as the medical history of the dataset included in the analysis are shown in **Table 2**. Of the participants imaged, 59.7% are female, 87.5% are white, and received 17.3 years of education (16 = college graduate; 18 = master’s degree). Less than 5% of them have high blood pressure, heart murmur, and anxiety disorders. Participants completed their 3T scan at a mean age of 45.7 years, approximately five years prior to the 7T scan (50.9 years).

**Table 2.** Demographics and medical history of 352 participants. SD stands for standard deviation.

|  | 3T | 7T |
| --- | --- | --- |
| Sample size, n | 352 |  |
| Sex female, n (%) | 210 (59.7) |  |
| Race white, n (%) | 308 (87.5) |  |
| Years of education, mean (SD) | 17.3 (2.7) |  |
| High blood pressure, n (%) | 5 (1.4) |  |
| Heart murmur, n (%) | 10 (2.8) |  |
| Heart surgery, n (%) | 0 |  |
| Diabetes, n (%) | 2 (0.6) |  |
| Depression, n (%) | 1 (0.3) |  |
| Panic attacks, n (%) | 3 (0.9) |  |
| Other anxiety disorder, n (%) | 12 (3.4) |  |
| Post traumatic disorder, n (%) | 0 |  |
| Age at scan, mean (SD) | 45.7 (9.2) | 50.9 (9.3) |
| Intracranial volume mm<sup>3</sup>, mean (SD) | 1.5521e+06 (1.4695e+05) | 1.4912e+06 (1.3767e+05) |

The quality control process also identified 6 cortical regions (entorhinal, parahippocampal, rostral anterior cingulate, caudal anterior cingulate, insula, and transverse temporal) where FreeSurfer was not able to generate accurate and consistent segmentations due to the presence of blood vessels and dura. Examples can be found in **Supplementary Figure S1**. We therefore removed these regions when comparing the correlation results between 3T and 7T. Statistics of the removed regions are still included in **Supplementary Table S1**. The correlation results therefore included 50 cortical volumes and 50 cortical thicknesses, and 38 subcortical volumes.

### Correlations of brain volumes and cortical thicknesses with age

Correlation between regional brain morphometrics and age from 352 pairs of 3T and 7T scans were calculated. **Figure 1** provides an overview of the results by categorizing the regions into total cortical grey matter volumes, total subcortical grey matter volumes, cerebral white matter volumes, and mean cortical thickness. For cortical and subcortical grey matter volumes, we saw both types of ICV corrections improved the correlation coefficient with age at 7T while weakening it at 3T. Pearson’s R values at 7T were significantly higher than at 3T using both the Residual and Proportional methods for both the cortical (pResidual < 1e-4, pProportional < 1e-7) and subcortical (pResidual < 1e-5, pProportional < 1e-3) grey matter volumes. While not notably changing the outcomes at 7T, the Residual method showed better R values at 3T when compared to the Proportional method. These results were independent of the ICV estimations, since they were almost identical at 3T and 7T (R=0.98, Figure 4). White matter volume showed weaker correlation with age than grey matter and had failed to show significance at 3T with both ICV correction methods. Mean cortical thickness which included sex only correction also failed to show significance at 3T while showing strong significance (p < 1e-10) and R value (-0.34) at 7T.

**Figure 1.**
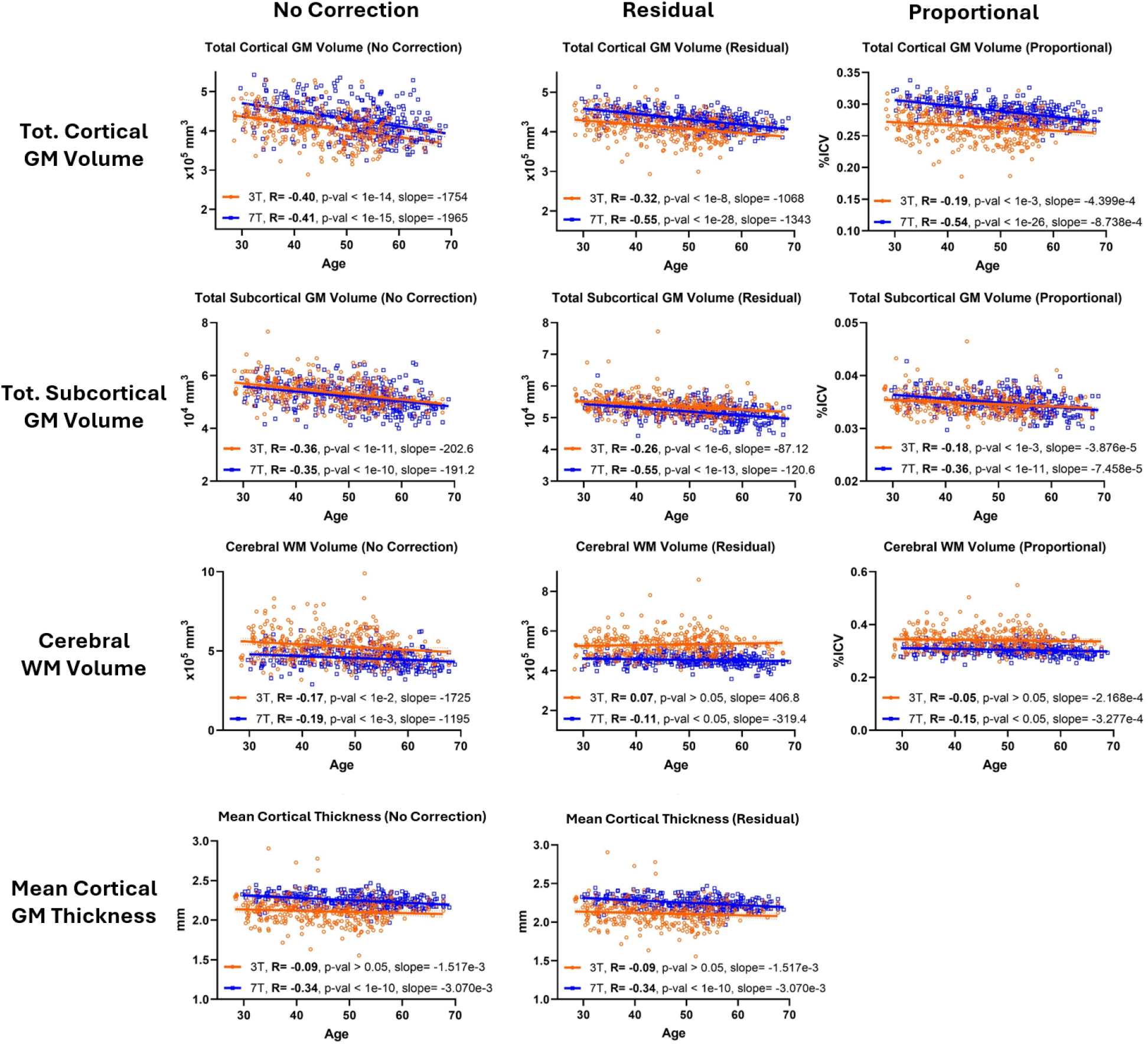
7T has a stronger inverse correlation of total cortical grey matter volume, total subcortical grey matter volume, total white matter volume, and mean cortical thickness with age. Brain morphometric correlations with age using 352 pairs of 3T and 7T MPRAGE scans, including the raw volumes (No Correction) and corrected for ICV using either residuals from regression model (Residual method) or division by ICV (Proportional method). Correlation between the ICV derived from 3T and 7T was shown to demonstrate consistent brain stripping results.

Pearson’s R values mapped into individual cortical regions are illustrated in **Figure 2**. The regions in the frontal and occipital lobe showed a generally stronger correlation than the temporal and parietal lobe. Regions with a strong correlation such as the superior frontal gyrus can be found significant at both 3T and 7T. **Supplementary Table S1** lists the correlation results of all regional brain volumes and cortical thicknesses after FDR correction (using the residual methods for ICV correction), as well as the respective linear regression slope and second-degree coefficient of the polynomial fit. When using the Residual method, 48 (50) out of 54 cortical volumes, 12 (40) out of 54 cortical thicknesses, and 25 (27) out of 38 sub-cortical volumes were found significantly correlated with age at 3T (7T). When using the Proportional method, 32 (53) out of 54 cortical volumes, 12 (40) out of 54 cortical thicknesses, and 21 (24) out of 38 sub-cortical volumes were found significant at 3T (7T).

**Figure 2.**
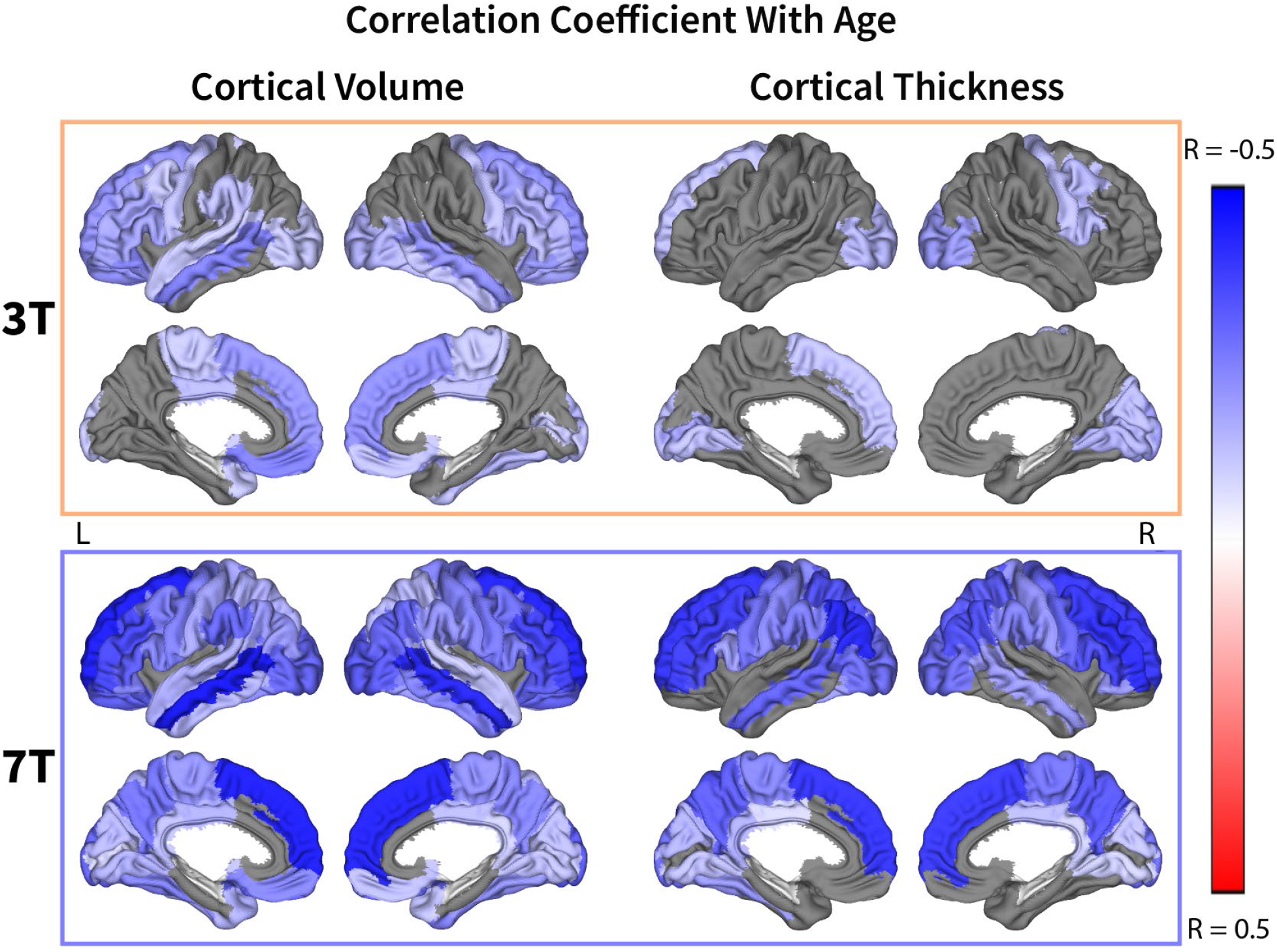
7T shows more significant regions in correlation of total grey matter volume or mean cortical thickness with age. Cortical regions corrected for ICV using Residual method and sex and found significant after FDR correction are shown with their respective Pearson correlation coefficient (positive correlation, blue; insignificant, grey; inverse correlation, red). Vessel-affected regions were removed. Cortical thicknesses were only corrected for sex.

The relationship between number of significant regions and sample size (N from 3 to 352) at 3T and 7T considering the different methods of ICV corrections is shown in **Figure 3**. For all brain volumes and cortical thicknesses combined, 32% (n = 111) of the 7T sample size were required to reach 85 significant regions found from the full 3T sample size (n = 352), corrected for ICV using the Residual method which achieves better correlation than the Proportional method most especially at 3T. When considering only cortical volumes (thicknesses), 73% (12%) of the 7T sample size were required to reach the same number of significant regions when compared with the full sample of 3T. For all 74 regions that were significant at both 3T and 7T, we compared the correlation coefficients and found that 20 (1, Optic Chiasm) regions had statistically stronger R value at 7T (3T) than the other field strength. **Supplementary Table S1** lists the p value for comparison between all regions.

**Figure 3.**
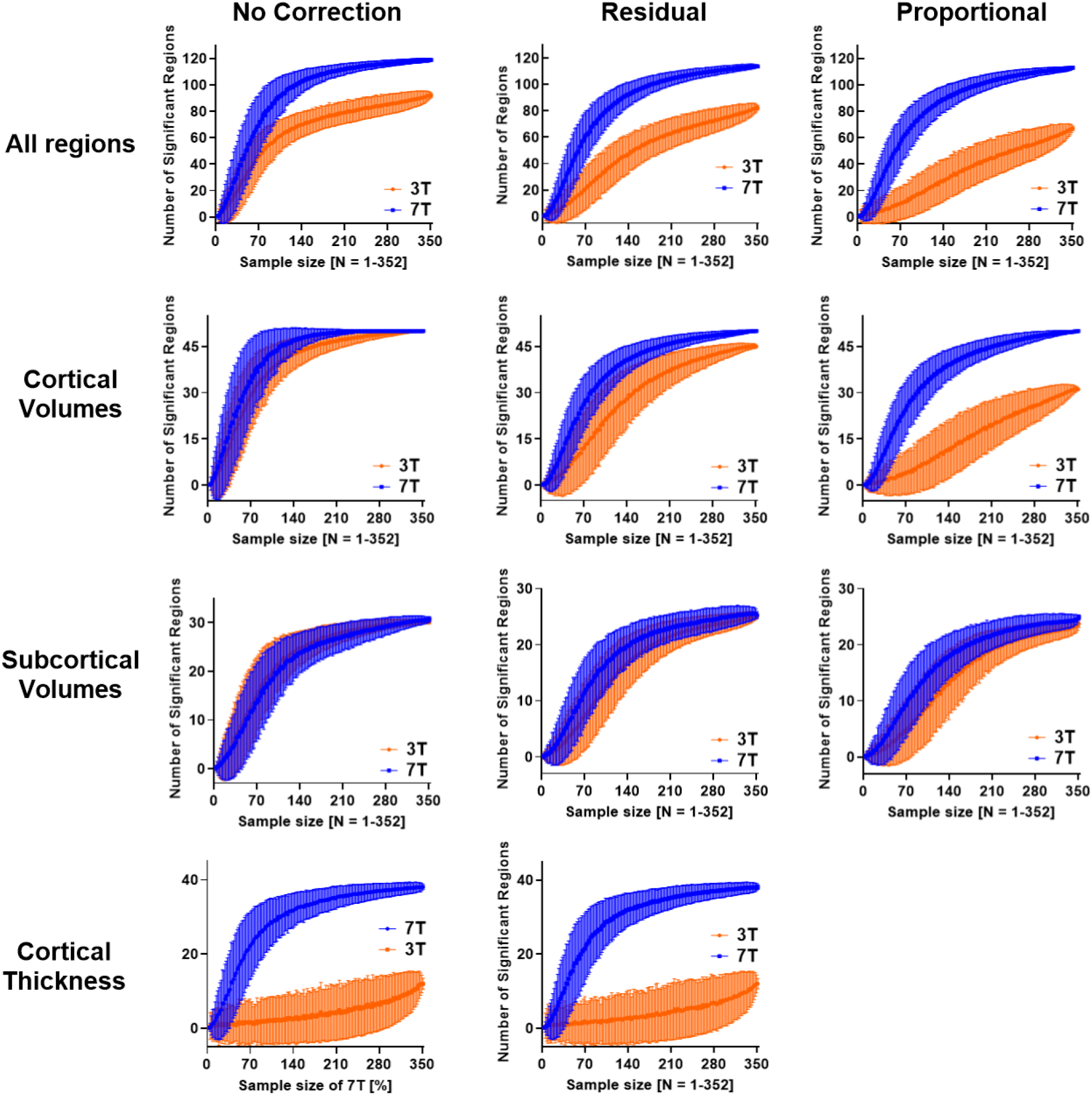
7T reduces required sample size in all regions, cortical volumes, subcortical volumes, and cortical thickness. Number of significant regions in raw volumes (no correction) and corrected for ICV using both the Residual and Proportional methods observed with increasing sample size significantly differed between 3T and 7T.

Figure 4. first affirmed the consistent ICV calculation between 3T and 7T, with a Pearson’s R=0.98. It then displayed the effect of ICV correction by showing the linear regression between corrected total cortical grey matter volume and ICV. Despite the fact that the Residual method proved to be effective at both 3T and 7T, interestingly we saw that the 7T images are less sensitive to the method of ICV correction.

We also mapped the cortical volume annual rate of change onto a brain atlas in **Figure 5**. The mean rate of change among regions with significant correlation with age was 0.32% for both 3T and 7T. We saw the frontal and occipital lobe, along with part of the temporal lobe volume decrease faster in general. The Residual method was used for this analysis.

**Figure 4.**
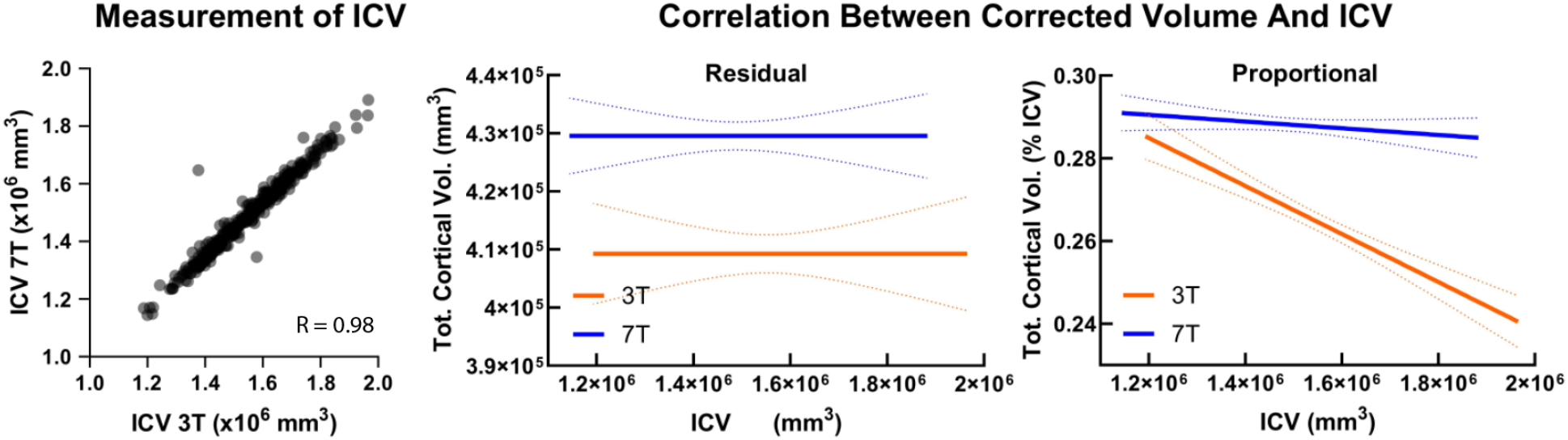
7T-derived ICV is consistent with that derived from 3T but more accurate in regional volumes. Comparison between the ICV value calculated at 3T and 7T as well as the effect of different ICV correction (Residual and Proportional) methods. Ideal correction should result in no correlation between total cortical volume and ICV. Dashed lines represent 95% confidence intervals. For the Residual method, both correlations showed no significant non-zero slope. For the Proportional method, 7T data showed no significant non-zero slope (p = 0.17) while the 3T data showed non-zero slope (p < 0.0001).

**Figure 5.**
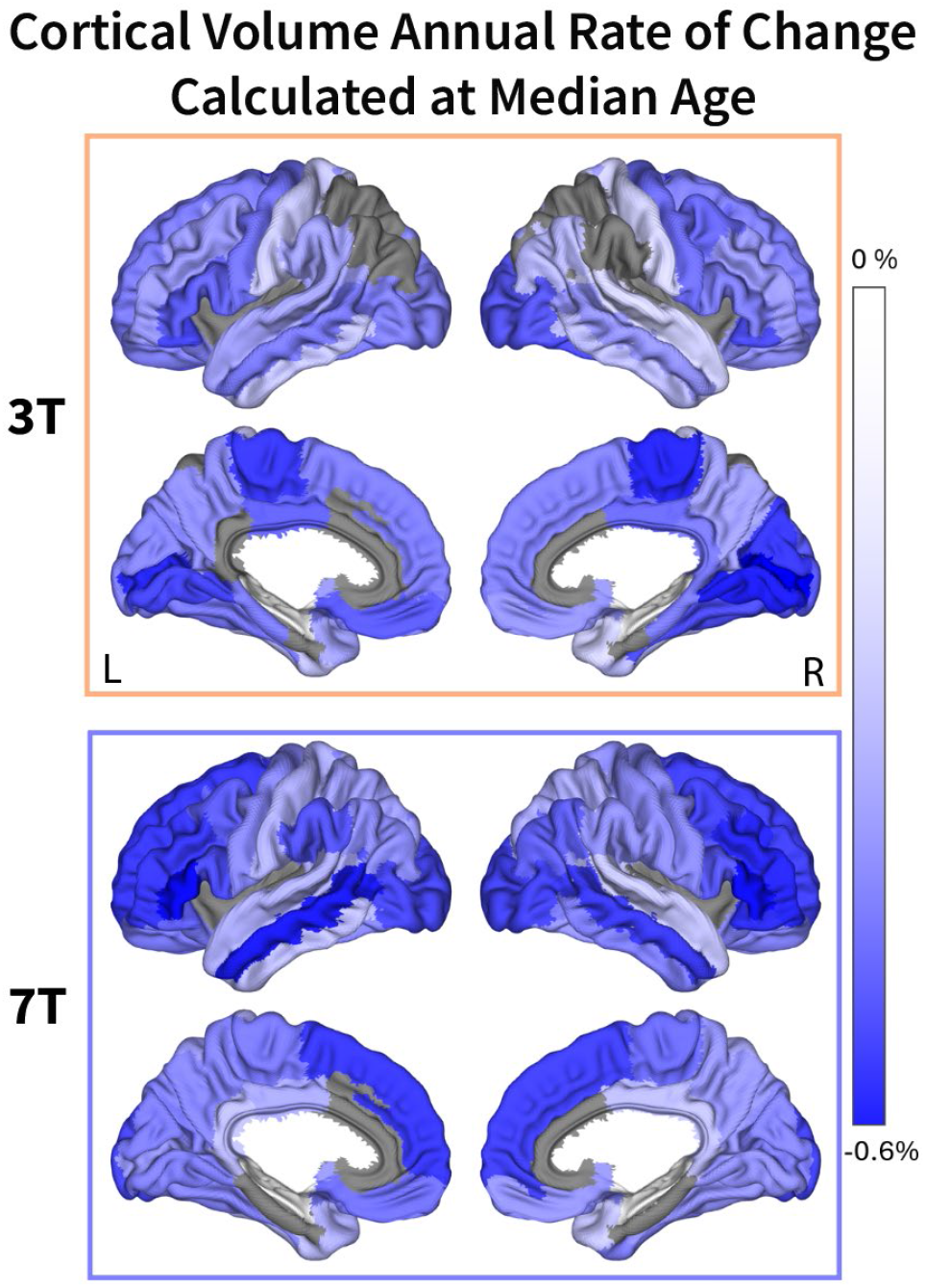
Mean cortical volume annual rate of change measured at 0.32% for both 3T and 7T. Cortical volumes were corrected for ICV using the Residual method, and the median age of the population used for this analysis is 52 years old.

## Discussion

In this study, we analyzed a large dataset of paired 3T and 7T MR images acquired on normal aging adults from 29 to 68 years of age, which allowed us to characterize the difference in their statistical performance when assessing cross-sectionally the brain morphometrics correlations as we age. We showed a heterogenous negative correlation between brain volume and age while providing an overview of how correlation of individual brain regions may be observed differently at either 3T or 7T. When subjected to a feasible scan time, less subjects would be necessary by scanning at 7T, providing studies with more flexibility to acquire additional sequences and/or save costs. When considering cortical thicknesses, which is required for AD cortical signature detection (Dickerson et al., 2009), only 7T provides sufficient brain coverage and sensitivity to aging effects.

The innovative radiofrequency coil developments (Andrea N Sajewski, 2023; Kim et al., 2016; Krishnamurthy et al., 2019; Santini et al., 2021; Santini et al., 2018) mitigated the excitation inhomogeneity traditionally observed at 7T MRI, potentially allowing more brain regions to be reliably quantified. Tailored preprocessing steps along with manual quality assurance and correction could further refine the automatic cortex parcellation by FreeSurfer.

The method of ICV correction greatly affects the results of correlation analysis, most notably in the 3T dataset. Previous studies (Wang et al., 2024) had investigated such effect on the correlation between brain volumetric measurements and cognitive performance at 3T, in which they showed that the regression based method was preferable in relation to the proportional method, which generated results that were biologically implausible. In our analysis, limited to morphometric variables alone, both correction methods were able to deliver plausible results, i.e., negative correlation between age and brain regional volumes. However, we noted that after correction using the Proportional method, the 3T dataset maintained a strong correlation between brain volumes and ICV, which could be interacting with the effect of age and thus reducing Pearson’s R values. The 7T dataset, on the other hand, showed little sensitivity as to which correction method was used. The Residual method seems to be the preferable one, since it gives its universal applicability.

Previous longitudinal studies (Fjell et al., 2013; Otsuka et al., 2022) on the relationship between grey matter volume and age reported an annual rate of change around 0.4% across brain regions. In our cross-sectional linear regression analysis, we derived a mean annual rate of change of 0.32% at both 3T and 7T at the study median age. The demographics as well as the image acquisition methods varied between our dataset and those of the published data. The Fjell study was using 1.5T MRI while the cohort lacked control for cognitive performance. The cross-sectional nature of our dataset may also play a role in the difference.

The white matter volume showed stability over aging at 3T and a small effect size, but significant, at 7T after adjusting for ICV. Studies have shown that white matter volume progresses differently with age compared to grey matter volume. Previous studies also showed modest changes in white matter volume until 40-50 years old before an accelerated decline (Fjell et al., 2014; Fjell, 2010). Hence a simple linear regression may not be enough to fully characterize the change of white matter volume with age. Further efforts shall be made to model the white matter volume by separating the age range while controlling other factors affecting white matter such as white matter hyperintensities and perivascular spaces (Fjell et al., 2014; Fjell, 2010).

As we gathered this large dataset of same-subject 3T and 7T scans, there was an average scan interval of 5.2 ± 4.5 years (after quality control). While the lack of field strength specific harmonization methods limited our ability to perform longitudinal analysis, the distance between the 3T and 7T scan resulted in different age distribution between groups. Fortunately, the quadratic terms are minimal in most regions, indicating that the relationship is nearly linear for the age range in this study and the impact of age differences at the scanning is relatively minor. At the moment of this analysis, information regarding comorbidities, lifestyle, cognition, AD risk factors, and other relevant factors presented in Table 2 were not included as covariates in the analysis, which could influence the change trajectory of brain morphometrics and will be subject of future study.

Our study also identified some limitations regarding the acquired datasets at both 3T and 7T. Firstly, FreeSurfer segmentations could fail when the cortex, occipital lobe specifically, had insufficient grey matter to white matter contrast sometimes observed in the 3T datasets. Control points were placed to aid the re-run of FreeSurfer segmentation. In the case of poor global contrast caused in conjunction with motion artifact, the subject is excluded. Cortical thickness measurements could also be biased due to the inconsistent tissue boundary due to the lack of white to gray matter contrast. Secondly, due to the altered T1 relaxation time, blood vessels, otherwise invisible at 3T, are marked with ultra bright contrast on 7T MPRAGE images. These blood vessels distinguish themselves with the surrounding tissue drastically, creating challenges for FreeSurfer algorithms which are tailored to lower field strengths image contrast. Major blood vessels such as the middle cerebral artery and pericallosal artery, were oftentimes included in the segmentation of their surrounding brain regions such as the anterior cingulate cortex, the parahippocampal cortex, the entorhinal cortex, the insula cortex, and the transverse temporal cortex in the 7T image segmentations. These regions were found to be either insignificant or having a weak and inconsistent correlation with age, which could be explained by inaccurate segmentation. We have excluded the most effected regions from this analysis. Another tissue that was more visible at 7T was the dura. While deep learning-based brain extraction provided consistent results across magnet strength and scanning procedure for our dataset, the survival rate of dura after brain stripping remained inconsistent, resulting in inconsistent over-classification of surface temporal cortical regions in the 7T segmentations. The effect of dura is manifested in the positive correlation between parahippocampal, entorhinal cortex thickness and age. The worsened susceptibility effects at 7T also gave rise to the air-tissue interface distortion artifact near the sinus in about half of the subjects, mainly presented in the inferior border of the orbital front cortex with extreme hyperintensity, veiling the cortex’s true boundary. While the impact to the volume measurement was limited, the susceptibility artifact appeared as an inevitable obstacle in calculating the true morphometrics of the region. Regarding the drawbacks encountered when segmenting the entorhinal and parahippocampal cortex with FreeSurfer, efforts have been made to address this issue: for instance, with the ASHS package and ASHS-PMC-T1 atlas, it is potentially possible to distinguish the complex tissues around the region and to generate more precise cortical/subcortical segmentations of the middle temporal lobe regions (Xie et al., 2016; Yushkevich et al., 2015).

## Conclusion

In this cohort of 352 participants with paired 3T and 7T scans, we compared the statistical performance in assessing age-related brain morphological changes. Our study reaffirmed the inverse correlation between brain volumes and cortical thicknesses and age and highlighted varying correlations in different brain regions at 3T and 7T. Compared to 3T, 7T has stronger correlations of total grey matter, subcortical, and white matter volumes, and mean cortical thickness with age, and shows more brain regions in which they volumes and cortical thicknesses have statistically significant correlations with age. For comparable statistical power at 3T, the required sample size for 7T is reduced for cortical and subcortical volumes, and substantially reduced for cortical thickness.

## Data Availability

All data produced in the present study are available upon reasonable request to the authors

## Acknowledgements

This research was supported by the National Institutes of Health (NIH-R56AG074467, NIH-R01AG053504, NIH-P01AG025204, NIH-RF1AG053504, NIH-R01MH111265, NIH-P01HL040962, and NIH-R01DK110041) and in part by the University of Pittsburgh Center for Research Computing, RRID:SCR_022735, through the resources provided. Specifically, this work used the HTC cluster, which is supported by NIH award number S10OD028483.

