## Supplemental Figure S1 for "Brain morphometrics correlations with age among 352 participants imaged with both 3T and 7T MRI: 7T improves statistical power and reduces required sample size"

**Figure S1.** Exclusion regions at 3T and 7T demonstrated inconsistent segmentation due to the presence of vessels and dura. While the dura could be seen on 3T, the vessels were completely invisible at 3T.

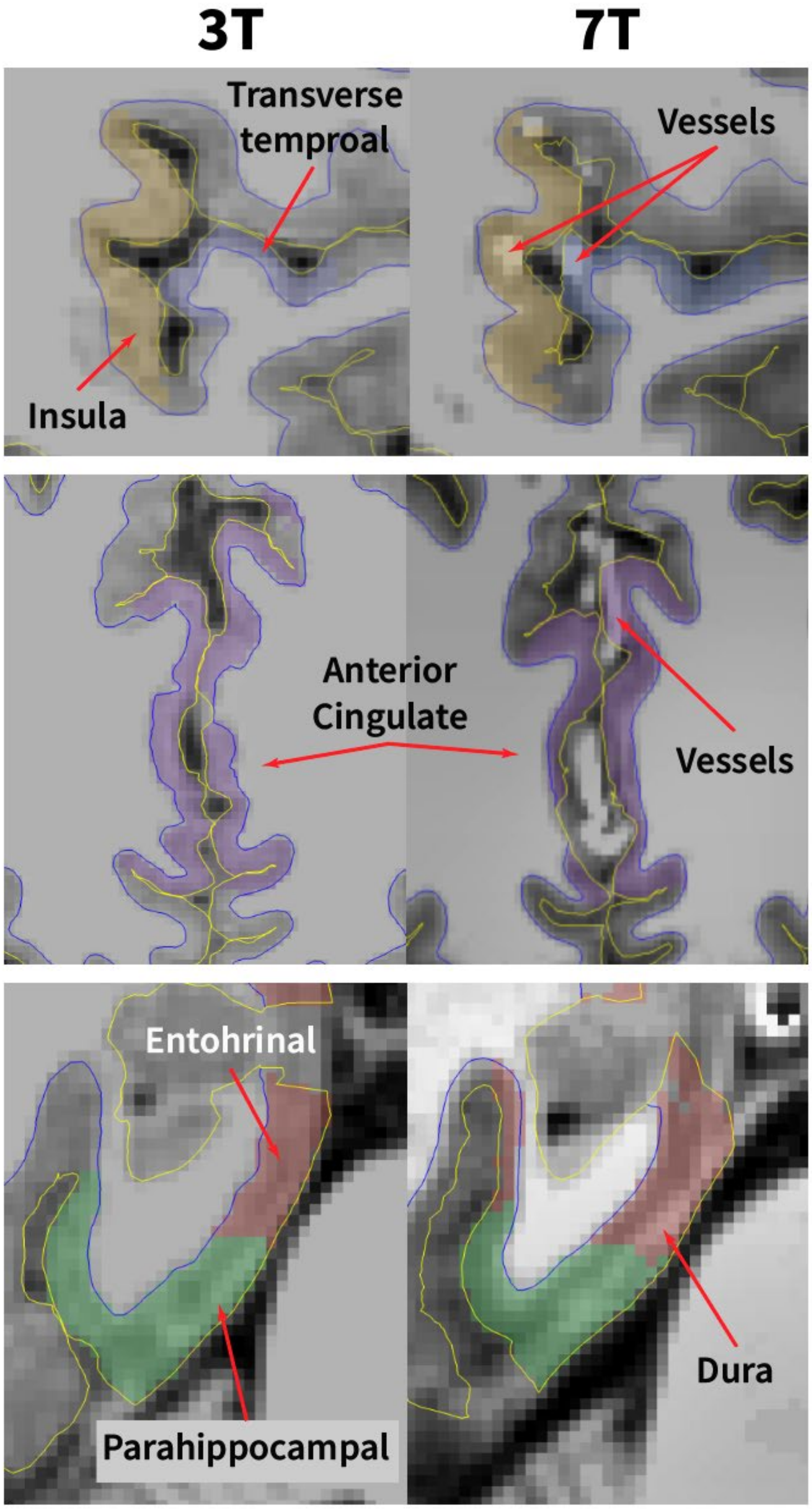
